# Estimating protection afforded by prior infection in preventing reinfection: Applying the test-negative study design

**DOI:** 10.1101/2022.01.02.22268622

**Authors:** Houssein H. Ayoub, Milan Tomy, Hiam Chemaitelly, Heba N. Altarawneh, Peter Coyle, Patrick Tang, Mohammad R. Hasan, Zaina Al Kanaani, Einas Al Kuwari, Adeel A. Butt, Andrew Jeremijenko, Anvar Hassan Kaleeckal, Ali Nizar Latif, Riyazuddin Mohammad Shaik, Gheyath K. Nasrallah, Fatiha M. Benslimane, Hebah A. Al Khatib, Hadi M. Yassine, Mohamed G. Al Kuwari, Hamad Eid Al Romaihi, Hanan F. Abdul-Rahim, Mohamed H. Al-Thani, Abdullatif Al Khal, Roberto Bertollini, Laith J. Abu-Raddad

## Abstract

**Background:** The Coronavirus Disease 2019 (COVID-19) pandemic has highlighted an urgent need to use infection testing databases to rapidly estimate effectiveness of prior infection in preventing reinfection (*PE*_*S*_) by novel variants of the severe acute respiratory syndrome coronavirus 2 (SARS-CoV-2).

**Methods:** Mathematical modeling was used to demonstrate the applicability of the test-negative, case-control study design to derive *PE*_*S*_. Modeling was also used to investigate effects of bias in *PE*_*S*_ estimation. The test-negative design was applied to national-level testing data in Qatar to estimate *PE*_*S*_ for SARS-CoV-2 infection and to validate this design.

**Results:** Apart from the very early phase of an epidemic, the difference between the test-negative estimate for *PE*_*S*_ and the true value of *PE*_*S*_ was minimal and became negligible as the epidemic progressed. The test-negative design provided robust estimation of *PE*_*S*_ even when *PE*_*S*_ began to wane after prior infection. Assuming that only 25% of prior infections are documented, misclassification of prior infection status underestimated *PE*_*S*_, but the underestimate was considerable only when >50% of the population was ever infected. Misclassification of latent infection, misclassification of current active infection, and scale-up of vaccination all resulted in negligible bias in estimated *PE*_*S*_. *PE*_*S*_ against SARS-CoV-2 Alpha and Beta variants was estimated at 97.0% (95% CI: 93.6-98.6) and 85.5% (95% CI: 82.4-88.1), respectively. These estimates were validated using a cohort study design.

**Conclusions:** The test-negative design offers a feasible, robust method to estimate protection from prior infection in preventing reinfection.

## Introduction

Estimating effectiveness of prior infection in preventing reinfection (*PE*_*S*_) is essential to understand the epidemiology of a given infection. Various studies have estimated *PE*_*S*_ for severe acute respiratory syndrome coronavirus 2 (SARS-CoV-2) variants.^1-9^However, there are challenges in estimating *PE*_*S*_ using conventional epidemiologic study designs. Such designs require extensive, complete electronic health records to be feasible. Vaccination scale-up makes it difficult to disentangle immunity induced by prior infection from that induced by vaccination. Even when it is feasible to apply conventional designs, estimates can be prone to strong bias, due to misclassification of prior infection status, since many prior infections are not documented.^10-13^ Effects of this bias increase with increased cumulative infection exposure in the population.^14^ Recent emergence of the Omicron^15^ (B.1.1.529) variant emphasized the need to estimate *PE*_*S*_ immediately for this variant.

Here, we demonstrate a robust, practical method to estimate *PE*_*S*_ using a test-negative, case-control study design. This is, to our knowledge, the first use of this method to estimate *PE*_*S*_. While it has been used to study vaccine effectiveness,^16-23^ it does not appear to have been used to

Estimate *PE*_*S*_, perhaps because of a perception that it is not applicable, as most prior and current infections are undocumented, unlike vaccinations, which are typically documented and tracked in health systems. We also validate this method by actually estimating *PE*_*S*_ for SARS-CoV-2 infection in Qatar, at a time when the Alpha^15^ (B.1.1.7) and Beta^15^ (B.1.351) variants dominated incidence.^22-27^

## Methods

### Test-negative case-control study design

The test-negative, case-control study design has emerged as a robust and practical method to assess vaccine effectiveness for respiratory tract infections.^16-23^ In this design, persons seeking healthcare because of symptoms are recruited into the study.^16, 17^ Those testing positive for the infection (cases) are then matched to those testing negative (controls).^16, 17^ Matching is done to control for differences in the risk of exposure to the infection.^22, 23, 28^ Vaccine effectiveness is then derived as one minus the ratio of the odds of vaccination in subjects testing positive to the odds of vaccination in subjects testing negative.^16, 17^ A key strength of this design is removal of differences in healthcare-seeking behavior between vaccinated and unvaccinated persons, thereby minimizing bias.^16, 17^ Another strength is minimization of bias arising from misclassification of infection.^16, 17^

### Mathematical modeling and simulation of the test-negative design

Mathematical modeling was used to demonstrate the applicability of the test-negative, case-control study design for deriving effectiveness of prior infection in preventing reinfection (*PE*_*S*_), that is, the proportional reduction in susceptibility to infection among those with prior infection versus those without.^2^ Modeling was also used to investigate effects of biases on estimated *PE*_*S*_. While this demonstration was done for SARS-CoV-2 infection, the approach is generic and should be broadly applicable to a range of infections.

Several models were devised to simulate SARS-CoV-2 infection transmission in the population and to investigate applicability of the test-negative design. The models were informed by previously published models for SARS-CoV-2 infection.^10, 13, 29-34^ The first was the classic Susceptible Exposed Infectious Recovered (SEIR) model, but extended to allow for reinfections (Baseline Model; Figure 1A). This model was used to demonstrate applicability of the test-negative design and to investigate sources of bias.

**Figure 1.**
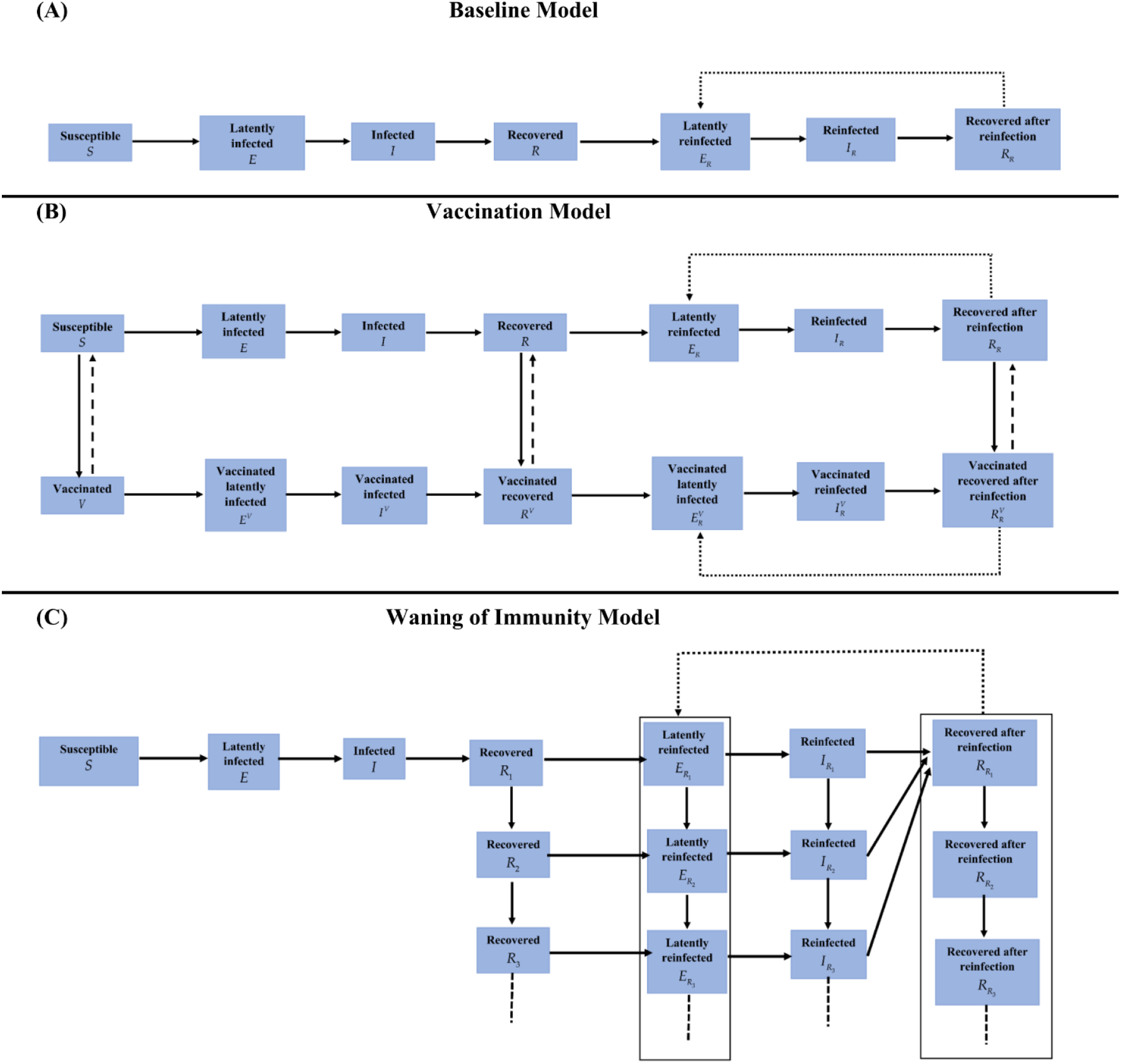
Schematic diagrams of mathematical models used in this study. A) Classic SEIR model extended to allow for reinfections (Baseline Model). B) Baseline Model extended to include vaccination (Vaccination Model). C) Baseline Model extended to include waning in protection of prior infection against reinfection (Waning of Immunity Model).

The second was an extension of the Baseline Model to incorporate scale-up of vaccination in the population (Vaccination Model; Figure 1B). This model was used to investigate whether vaccination could affect applicability of this method to estimate *PE*_*S*_. Vaccine effectiveness (*VE*_*S*_) was defined as the proportional reduction in susceptibility to infection among those vaccinated versus those unvaccinated.^33, 34^ *VE*_*S*_ was set at 75%, a representative value for the range of Coronavirus Disease 2019 (COVID-19) vaccines available at present. Duration of vaccine-induced protection was assumed to be 6 months in light of documented waning of COVID-19 vaccine protection.^26, 35^

The third was also an extension of the Baseline Model, incorporating gradual (linear) waning in protection offered by prior infection against reinfection (Waning of Immunity Model; Figure 1C).

These models consisted of coupled nonlinear differential equations that stratified the population into groups (compartments) based on infection status (infected, reinfected, or uninfected) and vaccination status (vaccinated, unvaccinated). Susceptible and recovered individuals (vaccinated or unvaccinated) were assumed at risk of acquiring the infection at a force of infection that varied throughout the epidemic due to variation in the contact rate.

These models were calibrated to mimic the actual evolution of the COVID-19 epidemic in Qatar.^13, 29^ The contact rate was varied to generate two major epidemic waves several months apart, as actually occurred.^13, 26, 29, 36^ Parameters of the models are summarized in Table 1. Further details on these models, their equations, and their parametrization can be found in previous publications.^10, 13, 29-34^ Modeling analyses were conducted in MATLAB R2019a (Boston/MA/USA).^37^

**Table 1.**
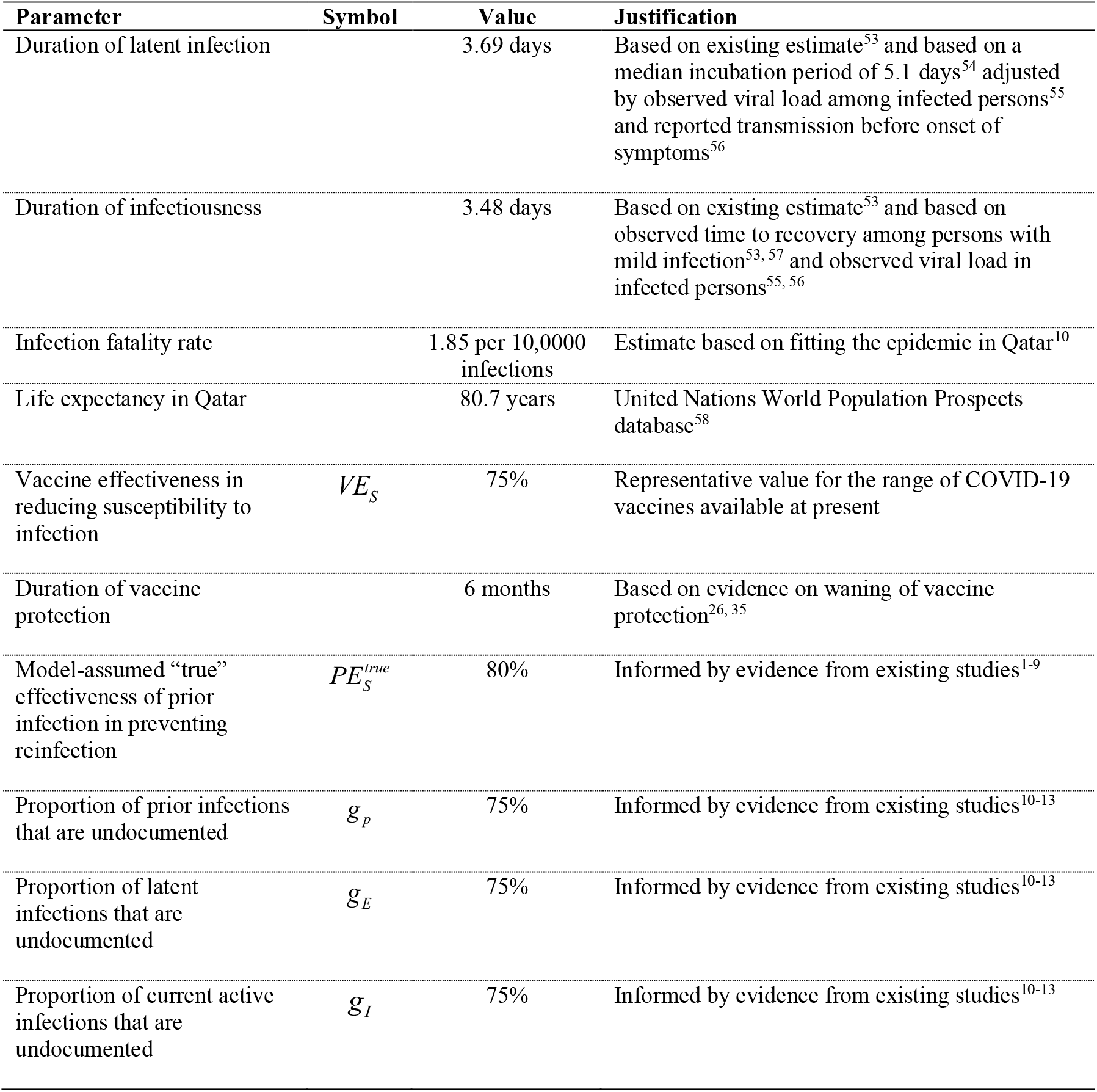
Model parameters and assumptions.

| Parameter | Symbol | Value | Justification |
| --- | --- | --- | --- |
| Duration of latent infection |  | 3.69 days | Based on existing estimate<sup>53</sup> and based on a median incubation period of 5.1 days<sup>54</sup> adjusted by observed viral load among infected persons<sup>55</sup> and reported transmission before onset of symptoms<sup>56</sup> |
| Duration of infectiousness |  | 3.48 days | Based on existing estimate<sup>53</sup> and based on observed time to recovery among persons with mild infection<sup>53, 57</sup> and observed viral load in infected persons<sup>55, 56</sup> |
| Infection fatality rate |  | 1.85 per 10,000 infections | Estimate based on fitting the epidemic in Qatar<sup>10</sup> |
| Life expectancy in Qatar |  | 80.7 years | United Nations World Population Prospects database<sup>58</sup> |
| Vaccine effectiveness in reducing susceptibility to infection | $VE_S$ | 75% | Representative value for the range of COVID-19 vaccines available at present |
| Duration of vaccine protection |  | 6 months | Based on evidence on waning of vaccine protection<sup>26, 35</sup> |
| Model-assumed “true” effectiveness of prior infection in preventing reinfection | $PE_S^{true}$ | 80% | Informed by evidence from existing studies<sup>1-9</sup> |
| Proportion of prior infections that are undocumented | $g_p$ | 75% | Informed by evidence from existing studies<sup>10-13</sup> |
| Proportion of latent infections that are undocumented | $g_E$ | 75% | Informed by evidence from existing studies<sup>10-13</sup> |
| Proportion of current active infections that are undocumented | $g_I$ | 75% | Informed by evidence from existing studies<sup>10-13</sup> |

### Effectiveness of prior infection against reinfection and impact of bias

Applying the test-negative, case-control study design, *PE*_*S*_ was derived as one minus the ratio of the odds of prior infection in subjects testing positive (such as by polymerase chain reaction (PCR) testing), to the odds of prior infection in subjects testing negative for the infection. The 2-by-2 table used to derive the odds ratio is shown in Figure 2A, as expressed in terms of the Baseline Model’s population variables. The mathematical expression for *PE*_*S*_ is also shown in Figure 2A, assuming no form of bias. An underlying assumption is that those being tested are a specific fixed proportion (sample) of all population variables.

**Figure 2.**
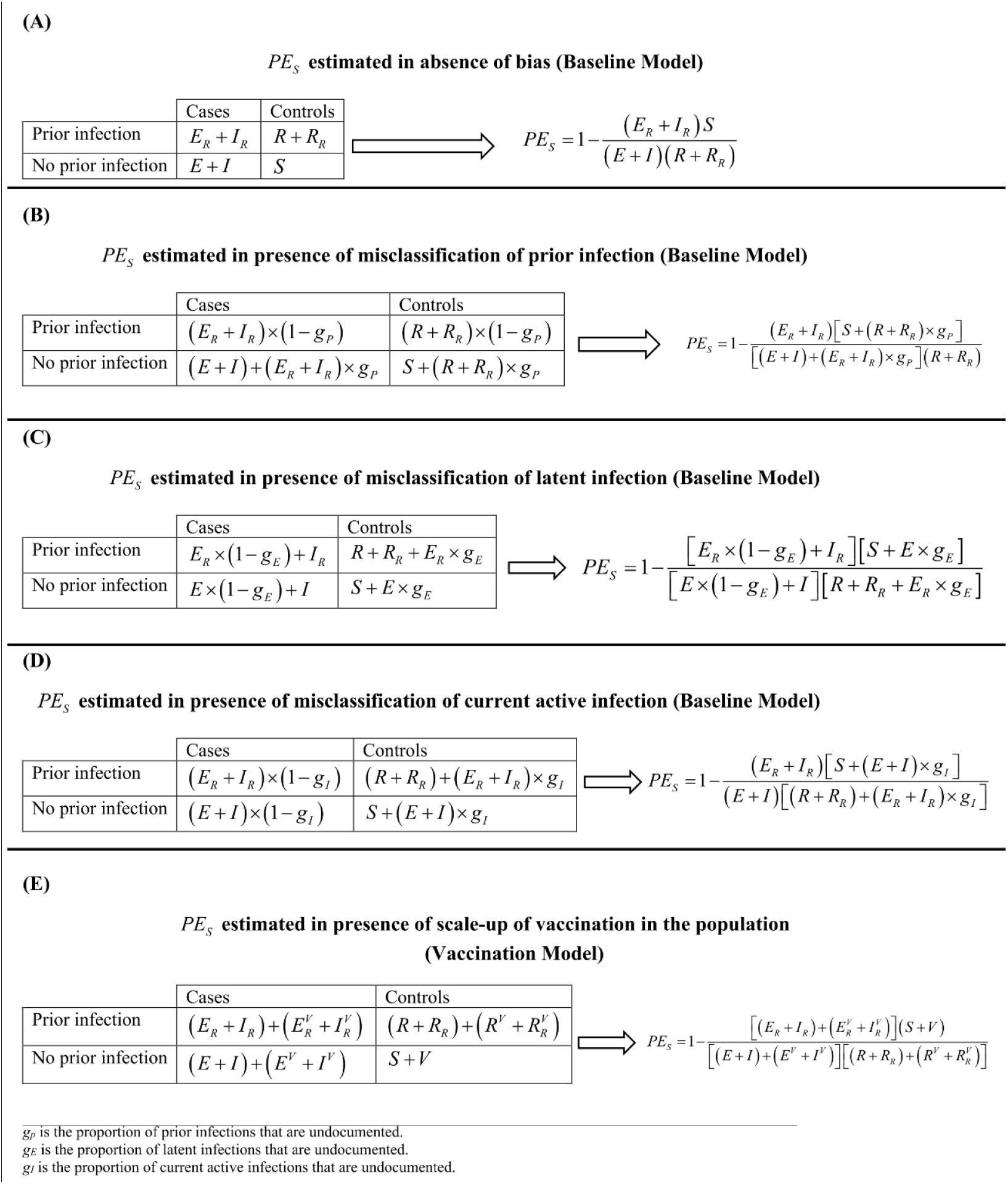
The 2-by-2 tables and equations used to estimate effectiveness of prior infection in preventing reinfection (*PE*_*S*_) using the test-negative, case-control study design. A) *PE*_*S*_ estimated in absence of bias. B) *PE*_*S*_ estimated in presence of misclassification of prior infection. C) *PE*_*S*_ estimated in presence of misclassification of latent infection. D) *PE*_*S*_ estimated in presence of misclassification of current active infection. E) *PE*_*S*_ estimated in presence of vaccination scale-up.

Several forms of bias may affect estimation of *PE*_*S*_ using this method. The most critical is misclassification of prior infection status. A proportion *g*_*P*_ of those previously infected may not have been diagnosed. They may have been unaware of their infections. It is reasonable to assume that most persons with a prior infection may not have had it documented.^10-13^ Here, we assumed that 75% of prior infections are undocumented (Table 1).

Unlike vaccine effectiveness studies, in which records are typically available to track vaccinations,^16-23, 28^ most persons with prior infection could be misclassified as persons with no prior infection. Similarly, most currently active infections may not be documented. The 2-by-2 table is thus modified for this bias along with the expression for *PE*_*S*_ (Figure 2B). It was assumed that this bias affects both cases and controls similarly, a valid assumption considering that both cases and controls are seeking healthcare because of symptoms. This assumption is central to the test-negative design strategy.^16, 17^

A second source of bias is misclassification of latent infection status. A proportion *g*_*E*_ of those with latent infections are asymptomatic; thereby remaining untested and undiagnosed. These cases would be misclassified as controls. The 2-by-2 table is thus modified to accommodate this bias along with the expression for *PE*_*S*_ (Figure 2C). We assumed that *g*_*E*_ = 75% (Table 1). We also assumed that this bias similarly affects those with and without prior infection. This is a valid assumption considering that both are seeking healthcare for the same reason, another assumption central to the test-negative design strategy.^16, 17^

A proportion *g*_*I*_ of cases (current active infections) could be misclassified as controls, because of lack of testing or due to imperfect sensitivity of the testing method, thereby introducing another form of bias. The 2-by-2 table is thus modified for this bias along with the expression for *PE*_*S*_ (Figure 2D). We assumed that *g*_*I*_ = 75% (Table 1). We also assumed that this bias similarly affects those with and without prior infection.^16, 17^

Estimation of *PE*_*S*_ may occur at a time when vaccination is being scaled up, as in the current COVID-19 pandemic. This could introduce bias as vaccination is another form of immune protection. Using the Vaccination Model, the 2-by-2 table is modified in presence of vaccination along with the expression for *PE*_*S*_ (Figure 2E). We assumed that vaccination is being linearly scaled up to reach the vaccine coverage attained in Qatar during the duration of the simulation. We also assumed that *PE*_*S*_ is the same regardless of vaccination status; that is, hybrid immunity of prior infection and vaccination is superior to that of either prior infection or vaccination separately.^36, 38^

Since different forms of bias may act synergistically when present together, the impact of the above biases was also investigated by applying all of them simultaneously.

### Real-world application: Effectiveness of prior infection in preventing reinfection in Qatar

To validate the test-negative design, *PE*_*S*_ was estimated in Qatar using national-level routine PCR testing data. Only persons being PCR tested for clinical suspicion of infection due to symptoms between March 8 and April 21, 2021 were eligible for inclusion in this analysis. This study duration was chosen because there are existing estimates for *PE*_*S*_ during this time, but using a conventional, cohort study design.^4^ This allows validation of the estimate generated using the test-negative design.

Prior infection was defined as a PCR-confirmed infection ≥90 days before a new PCR-positive test.^2, 6^ Individuals infected during the 90 days preceding the PCR test were thus excluded. Based on existing evidence^39-41^ and viral genome sequencing,^3, 22^ a SARS-CoV-2 Alpha variant case was defined as an S-gene “target failure” case using the TaqPath COVID-19 Combo Kit platform (Thermo Fisher Scientific, USA^42^) applying the criterion of a PCR cycle threshold (Ct) value ≤30 for both the N and ORF1ab genes, but a negative outcome for the S gene.^3, 4, 41^ With essentially only Beta and Alpha cases identified between March 8 and April 21, 2021,^22-27^ a Beta case was proxied as the complement of the Alpha criterion, that is, any case with a Ct value ≤30 for the N, ORF1ab, and S genes.

Only the first PCR-positive test during the study was included for each case, and only the first PCR-negative test during the study was included for each control, per established protocol for the test-negative design.^22, 23, 26, 28^ Alpha and Beta cases were exact matched one-to-one to controls (PCR-negative persons) by sex, 10-year age group, nationality, and calendar week of PCR test. Matching of cases and controls was done to control for known differences in the risk of exposure to SARS-CoV-2 infection in Qatar.^13, 43-46^ Further description of Qatar’s databases and methods of analysis can be found in previous publications.^1-4, 22, 23, 26, 28, 36, 43^

Socio-demographic characteristics of study samples were described using frequency distributions and measures of central tendency. The odds ratio, comparing odds of prior infection among cases versus controls, and its associated 95% confidence interval (CI) were derived using conditional logistic regression, factoring matching in the study design. This analytical approach is done to minimize potential bias due to variation in epidemic phase^16, 47^ and other confounders.^13, 43-46, 48,49^ *PE*_*S*_ and its associated 95% CI were calculated by applying the following equation:

*PE*_*S*_ = 1™ odds ratio of prior infection among cases versus controls

Statistical analyses were conducted in STATA/SE version 17.0.^50^ The study was approved by the Hamad Medical Corporation and Weill Cornell Medicine-Qatar Institutional Review Boards with waiver of informed consent.

## Results

### Protection of prior infection using the test-negative design and impact of bias

Figure 3 shows simulated evolution of the SARS-CoV-2 epidemic in its two waves (Figure 3A), the proportion of the population ever infected (Figure 3B), and vaccine coverage (Figure 3C). Figure 4A shows the estimated *PE*_*S*_ using the test-negative design (labeled as 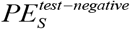), by application of the expression in Figure 2A, compared to the true *PE*_S_ (labeled as 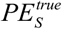), here assumed at 80% (Table 1). Apart from the very early phase of the epidemic, when the number of reinfections was minimal, the difference between 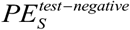 and 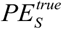 was no more than several percentage points. The difference became negligible as the epidemic progressed.

**Figure 3.**
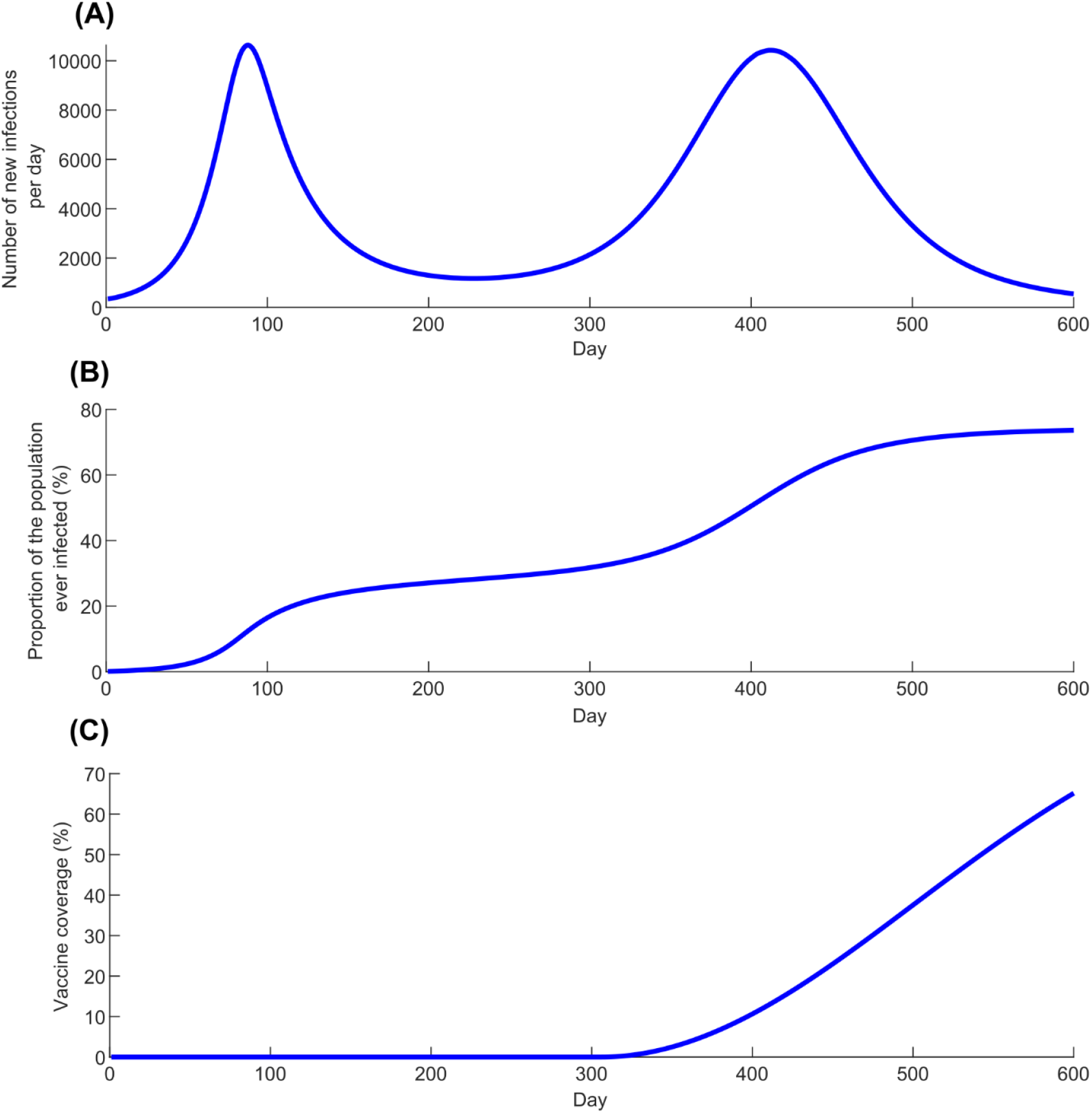
Simulated SARS-CoV-2 epidemic through two epidemic waves. A) Daily number of new infections. B) Proportion of the population ever infected. C) Scale-up of vaccine coverage.

**Figure 4.**
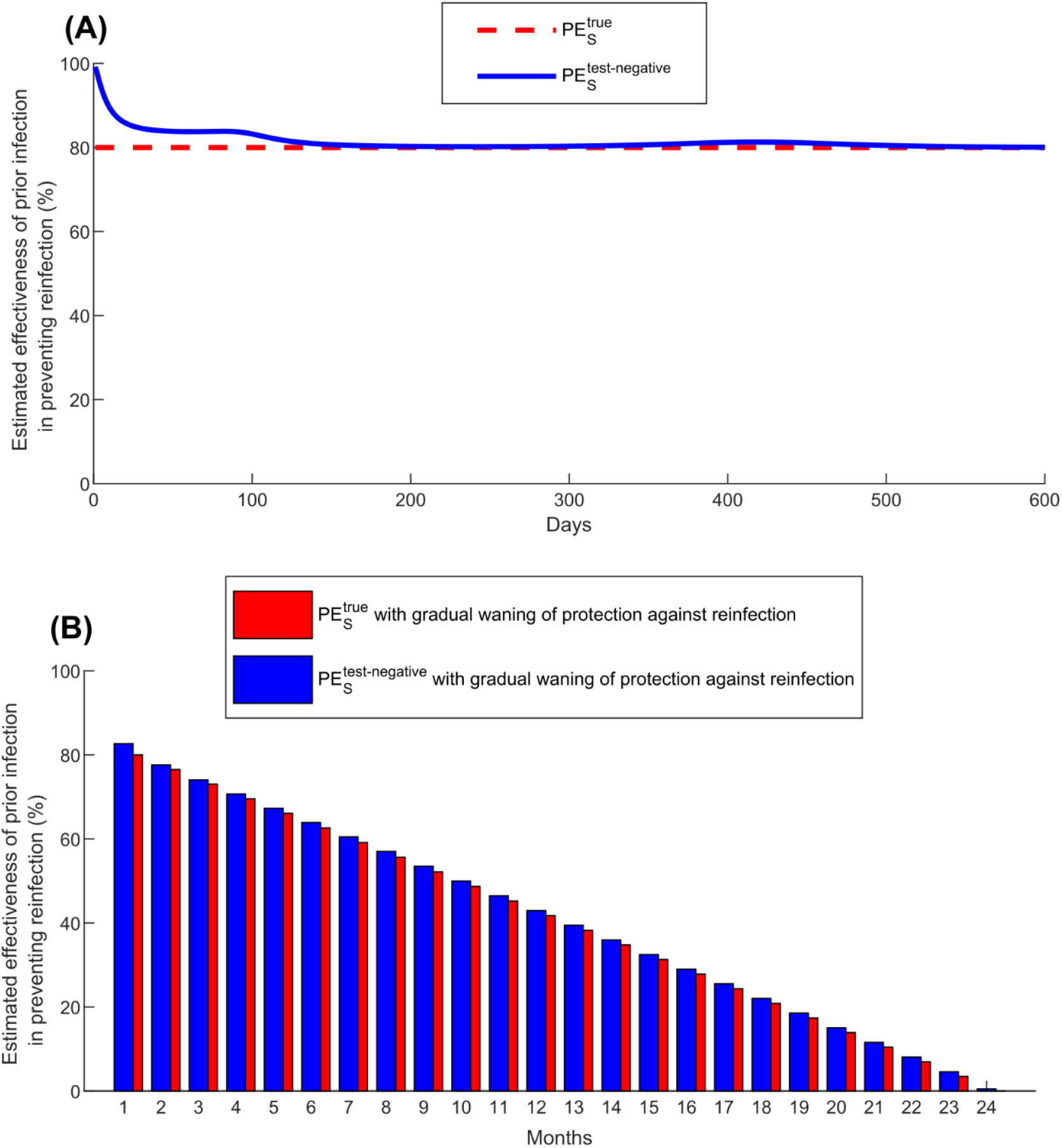
Estimated effectiveness of prior infection in preventing reinfection using the test-negative study design 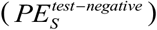 compared with the true effectiveness of prior infection in preventing reinfection 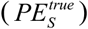. A) 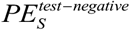 versus 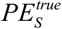 in presence of no waning of protection (Baseline Model). B) 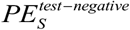 versus 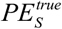 month by month after the prior infection in presence of gradual waning of protection against reinfection (Waning of Immunity Model).

Assuming that only 25% of prior infections are documented (Table 1), Figure 5A shows the impact of misclassification of prior infection, by application of the expression in Figure 2B. This form of bias resulted in underestimation of 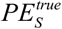. When the proportion of the population ever infected was below 50% (Figure 3B), 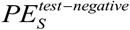 was only few percentage points lower than that of 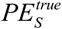. However, the underestimation increased to as much as 30 percentage points when the proportion of the population ever infected was ∼75%.

**Figure 5.**
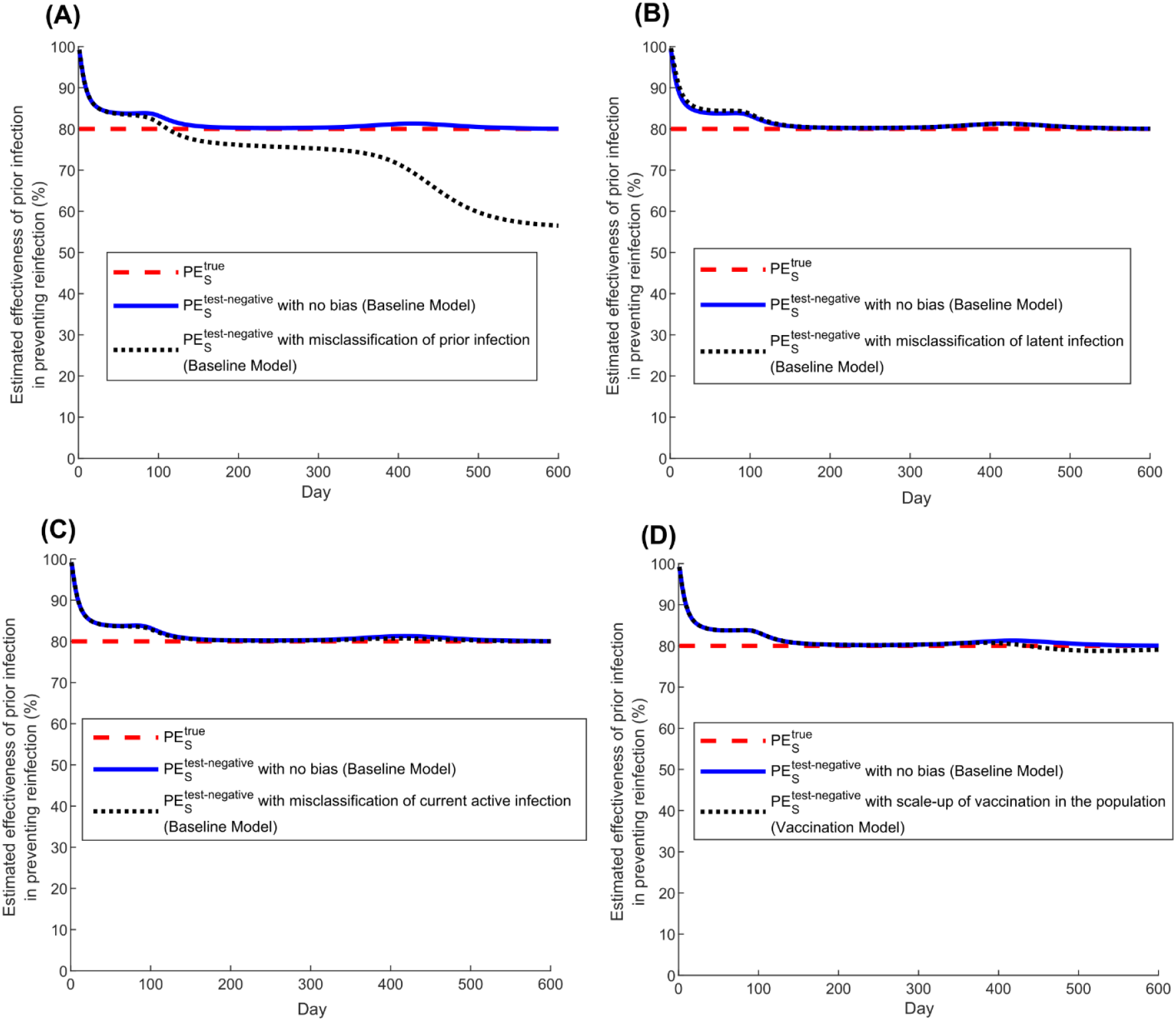
Impact of bias in estimating effectiveness of prior infection in preventing reinfection using the test-negative study design 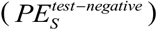). A) Impact of misclassification of prior infection. B) Impact of misclassification of latent infection. C) Impact of misclassification of current active infection. D) Impact of scale-up of vaccination in the population.

Misclassification of latent infection (Figure 5B), misclassification of current active infection (Figure 5C), and scale-up of vaccination (Figure 5D), all resulted in negligible bias in estimated 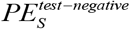. Application of the above forms of bias at the same time suggested that there is no synergy when biases are combined (Figure S1).

Applying the Waning of Immunity Model, Figure 4B shows 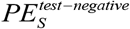 versus 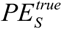, month by month after prior infection, assuming that there is a gradual linear waning in protection of prior infection against reinfection. This comparison was done after the second wave at day 600 after the virus introduction (Figure 3A). 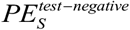 provided a robust approximation of 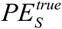 and its waning month by month.

### Application: Effectiveness of prior infection in preventing reinfection in Qatar

Figure 6 presents a flowchart describing the population selection process for estimating *PE*_*S*_ in Qatar using the test-negative design. The median age of study subjects was 32-34 years, at least half were males, and they came from diverse countries (Table 2). Study samples were broadly representative of Qatar’s demographics.^43, 51^

**Figure 6.**
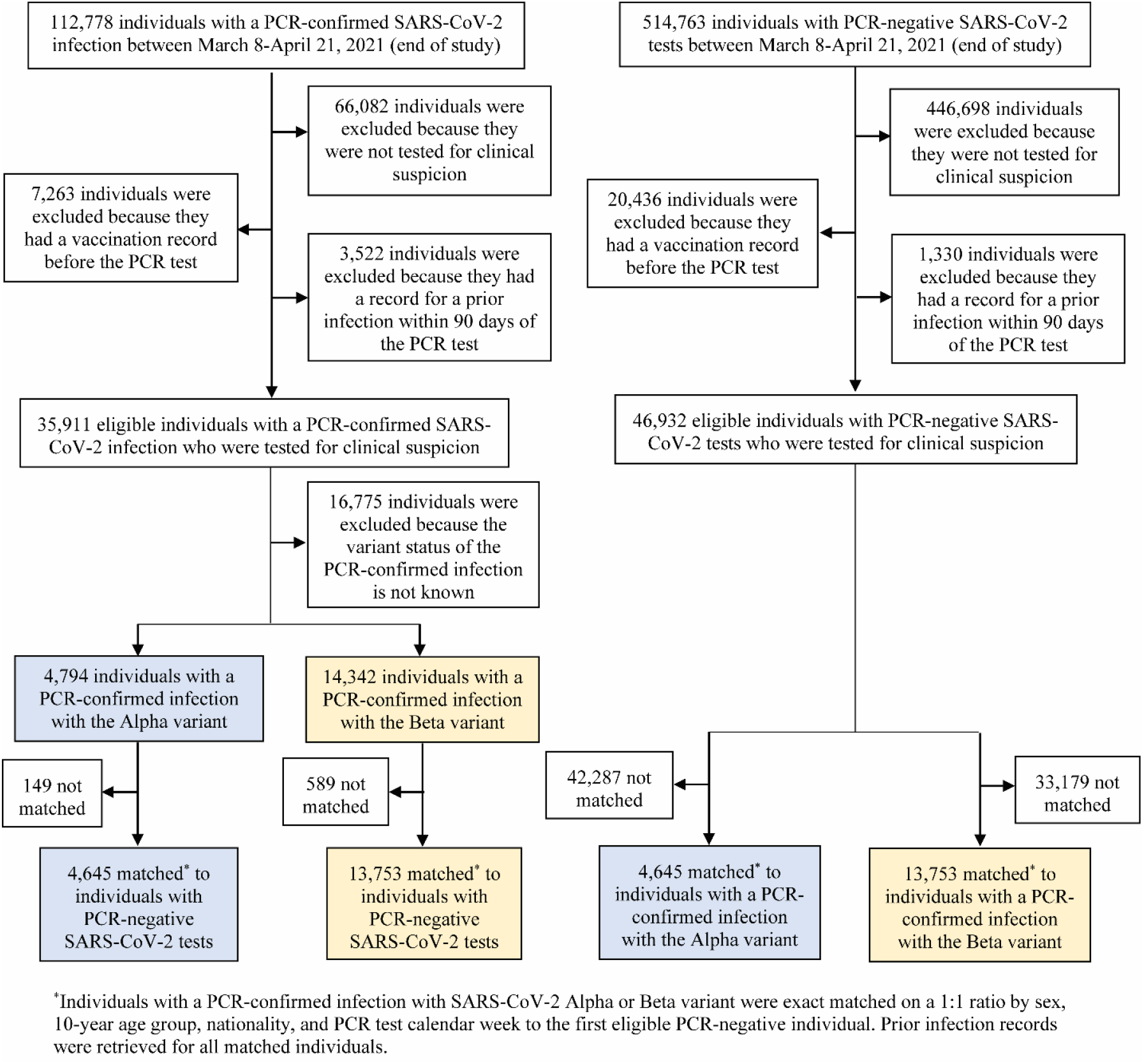
Flowchart describing the population selection process to estimate effectiveness of prior infection in preventing reinfection using the test-negative study design.

**Table 2.** Demographic characteristics of subjects in the samples used to estimate effectiveness of prior infection in preventing reinfection using the test-negative study design.

| Characteristics | Cases<sup>*</sup><br>(PCR-confirmed infection<br>with the Alpha variant) | Controls<sup>*</sup><br>(PCR-negative) | SMD<sup>§</sup> | Cases<sup>*</sup><br>(PCR-confirmed infection<br>with the Beta variant) | Controls<sup>*</sup><br>(PCR-negative) | SMD<sup>§</sup> |
| --- | --- | --- | --- | --- | --- | --- |
|  | N=4,645 | N=4,645 |  | N=13,753 | N=13,753 |  |
| **Median age (IQR) — years** | 33 (25-40) | 32 (24-40) | 0.01<sup>¶</sup> | 34 (27-40) | 33 (27-40) | 0.01<sup>¶</sup> |
| **Age group — no. (%)** |  |  |  |  |  |  |
| <20 years | 868 (18.7) | 868 (18.7) |  | 1,767 (12.9) | 1,767 (12.9) |  |
| 20-29 years | 923 (19.9) | 923 (19.9) |  | 2,931 (21.3) | 2,931 (21.3) |  |
| 30-39 years | 1648 (35.5) | 1648 (35.5) |  | 5,213 (37.9) | 5,213 (37.9) |  |
| 40-49 years | 871 (18.8) | 871 (18.8) | 0.00 | 2,877 (20.9) | 2,877 (20.9) | 0.00 |
| 50-59 years | 272 (5.9) | 272 (5.9) |  | 797 (5.8) | 797 (5.8) |  |
| 60-69 years | 53 (1.1) | 53 (1.1) |  | 132 (1.0) | 132 (1.0) |  |
| 70+ years | 10 (0.2) | 10 (0.2) |  | 36 (0.3) | 36 (0.3) |  |
| **Sex** |  |  |  |  |  |  |
| Male | 2,339 (50.4) | 2,339 (50.4) | 0.00 | 9,467 (68.8) | 9,467 (68.8) | 0.00 |
| Female | 2,306 (49.6) | 2,306 (49.6) |  | 4,286 (31.2) | 4,286 (31.2) |  |
| **Nationality<sup>†</sup>** |  |  |  |  |  |  |
| Bangladeshi | 235 (5.1) | 235 (5.1) |  | 1,334 (9.7) | 1,334 (9.7) |  |
| Egyptian | 358 (7.7) | 358 (7.7) |  | 990 (7.2) | 990 (7.2) |  |
| Filipino | 764 (16.5) | 764 (16.5) |  | 1,610 (11.7) | 1,610 (11.7) |  |
| Indian | 789 (17.0) | 789 (17.0) |  | 3,481 (25.3) | 3,481 (25.3) |  |
| Nepalese | 170 (3.7) | 170 (3.7) | 0.00 | 1,283 (9.3) | 1,283 (9.3) | 0.00 |
| Pakistani | 192 (4.1) | 192 (4.1) |  | 542 (3.9) | 542 (3.9) |  |
| Qatari | 762 (16.4) | 762 (16.4) |  | 1,288 (9.4) | 1,288 (9.4) |  |
| Sri Lankan | 125 (2.7) | 125 (2.7) |  | 538 (3.9) | 538 (3.9) |  |
| Sudanese | 166 (3.6) | 166 (3.6) |  | 442 (3.2) | 442 (3.2) |  |
| Other nationalities<sup>‡</sup> | 1,084 (23.3) | 1,084 (23.3) |  | 2,245 (16.3) | 2,245 (16.3) |  |

Abbreviations: IQR, interquartile range; PCR, polymerase chain reaction; SMD, standardized mean difference.
<sup>\*</sup>Cases and controls were matched one-to-one by sex, 10-year age group, nationality, and calendar week of PCR test.
<sup>†</sup>Nationalities were chosen to represent the most populous groups in Qatar.
<sup>‡</sup>These comprise 61 other nationalities in Qatar in the Alpha variant analysis and 78 other nationalities in the Beta variant analysis.
<sup>§</sup>SMD is the difference in the mean of a covariate between groups divided by the pooled standard deviation. An SMD<0.1 indicates adequate matching.
<sup>¶</sup>SMD is the mean difference between groups divided by the pooled standard deviation.

Among the 4,645 Alpha cases (PCR-positive persons), 7 had a record of prior infection, compared to 232 among their matched controls (PCR-negative persons). *PE*_*S*_ against Alpha was estimated at 97.0% (95% CI: 93.6-98.6). Among the 13,753 Beta cases, 124 had a record of prior infection, compared to 815 among their matched controls. *PE*_*S*_ against Beta was estimated at 85.5% (95% CI: 82.4-88.1).

During the study duration (March 8, 2021 to April 21, 2021), we conducted earlier two matched cohort studies to estimate *PE*_*S*_ for Alpha and for Beta.^4^ For Alpha, cohort-study estimates were 97.6% (95% CI: 95.7-98.7%) and 96.4% (95% CI: 92.1-98.3%).^4^ For Beta, cohort-study estimates were 92.3% (95% CI: 90.3-93.8%) and 86.4% (95% CI: 82.5-89.5%).^4^

## Discussion

Study results show that the test-negative design can be used to generate rigorous estimates for protection afforded by prior infection against reinfection, even though most prior infections are undocumented. Estimates were robust despite several forms of potential bias, and even under rather extreme assumptions for these biases. The test-negative design was also applied to Qatar’s routine PCR testing data, and results were validated by comparing test-negative estimates to those generated using conventional cohort study designs.^4^ Application of the test-negative design should be feasible in different countries as long as there are databases for infection testing that are of reasonable quality and that can be linked to documented prior infection status (and preferably to vaccination status). Such databases are available and have been used extensively in vaccine effectiveness studies, such as for SARS-CoV-2 infection.^18-23, 28^

Of the considered biases, only misclassification of prior infection status could have a large effect on *PE*_*S*_ estimation, but mainly where more than 50% of the population already had a prior infection. This situation is unlikely to have been reached for SARS-CoV-2 infection in most countries.^52^ Even in such situations, the direction (and magnitude) of bias is known; it underestimates *PE*_*S*_. Therefore, the test-negative design can still provide a lower bound for the true *PE*_*S*_, which may be sufficient to inform public health decision making. Thus, this bias may not restrict the utility of this method.

The test-negative study design has strengths that conventional designs may lack. Cohort study designs can be affected by bias resulting from different infection testing frequencies in the two arms of the study. This bias does not affect the test-negative design, as it uses only those who are tested. An example can be seen in comparing the results of the test-negative design to the results of our earlier cohort design.^4^ In the cohort design, adjustment for testing frequency reduced *PE*_*S*_ from 97.6% (95% CI: 95.7-98.7%) to 95.8% (95% CI: 92.5-97.7%) for Alpha,^4^ very similar to the test-negative estimate of 97.0% (95% CI: 93.6-98.6). Similarly for Beta, adjustment for testing frequency reduced *PE*_*S*_ from 92.3% (95% CI: 90.3-93.8%) to 86.5% (95% CI: 83.0-89.2%),^4^ very similar to the test-negative estimate of 85.5% (95% CI: 82.4-88.1). Accordingly, the test-negative design may provide a more robust estimate than the cohort design.

The test-negative design may also be preferable to the cohort design for other reasons. Cohort designs rely on cohorts that may not be strictly comparable, and it may not be possible to control for all differences in risk of exposure to the infection by matching and analysis adjustments. For example, in our earlier cohort study,^4^ we compared those with a record of a prior PCR-confirmed infection to those with an antibody-negative test, but these two groups may differ in ways that cannot be controlled. Meanwhile, the test-negative design is perhaps less susceptible to such differences, as cases and controls are intentionally recruited for the same healthcare-seeking behavior. Lastly, while the test-negative design can be biased by misclassification of prior infection, the cohort design is perhaps more affected by this bias. The odds ratio metric in the test-negative design is less affected by this bias than the relative risk, incidence rate ratio, or hazard ratio metrics in the cohort design.

In regard to limitations, specific forms of bias were investigated, but other sources of bias are possible, and these may depend on the database being analyzed.^26^ There is already a volume of literature investigating other forms of bias for the test-negative design in the context of vaccine effectiveness estimation,^16, 17^ some of which may apply in the context of *PE*_*S*_ estimation. While this study demonstrated use of the test-negative design to estimate *PE*_*S*_, other factors need to be considered in actual application. For instance, the algorithm for matching needs to be developed with knowledge of the local epidemiology to ensure that matching can effectively control differences in the risk of exposure to the infection.

In conclusion, the test-negative design offers a feasible and robust method to estimate protection of prior infection in preventing reinfection. This method should be considered to provide rapid, rigorous estimates of protection offered by prior infection for different variants of SARS-CoV-2, such as the Omicron that emerged recently.

## Data Availability

The dataset of this study is a property of the Qatar Ministry of Public Health that was provided to the researchers through a restricted-access agreement that prevents sharing the dataset with a third party or publicly. Future access to this dataset can be considered through a direct application for data access to Her Excellency the Minister of Public Health (https://www.moph.gov.qa/english/Pages/default.aspx). Aggregate data are available within the manuscript and its Supplementary information.

## Acknowledgements

We acknowledge the many dedicated individuals at Hamad Medical Corporation, the Ministry of Public Health, the Primary Health Care Corporation, the Qatar Biobank, Sidra Medicine, and Weill Cornell Medicine – Qatar for their diligent efforts and contributions to make this study possible.

HHA acknowledges the joint support of Qatar University and Marubeni M-QJRC-2020-5. The authors are grateful for support from the Biomedical Research Program and the Biostatistics, Epidemiology, and Biomathematics Research Core, both at Weill Cornell Medicine-Qatar, as well as for support provided by the Ministry of Public Health, Hamad Medical Corporation, and Sidra Medicine. The authors are also grateful for the Qatar Genome Programme and Qatar University Biomedical Research Center for institutional support for the reagents needed for the viral genome sequencing. Statements made herein are solely the responsibility of the authors. The funders of the study had no role in study design, data collection, data analysis, data interpretation, or writing of the article. The developed mathematical models were made possible thanks to modeling infrastructure developed through NPRP grant number 9-040-3-008 (Principal investigator: LJA) and NPRP grant number 12S-0216-190094 (Principal investigator: LJA) from the Qatar National Research Fund (a member of Qatar Foundation; https://www.qnrf.org). The statements made herein are solely the responsibility of the authors. The funders had no role in study design, data collection and analysis, decision to publish, or preparation of the manuscript.

## Author contributions

HHA and MT constructed, coded, and parameterized the mathematical models, conducted the mathematical modeling analyses, and co-wrote the first draft of the article. HC performed the statistical analyses and co-wrote the first draft of the article. LJA conceived and designed the study, led the mathematical modeling and statistical analyses, and co-wrote the first draft of the article. PT and MRH conducted the multiplex, RT-qPCR variant screening and viral genome sequencing. HY, FMB, and HAK conducted viral genome sequencing. All authors contributed to data collection and acquisition, database development, discussion and interpretation of the results, and to the writing of the manuscript. All authors have read and approved the final manuscript.

## Supplementary Material

**Figure S1.**
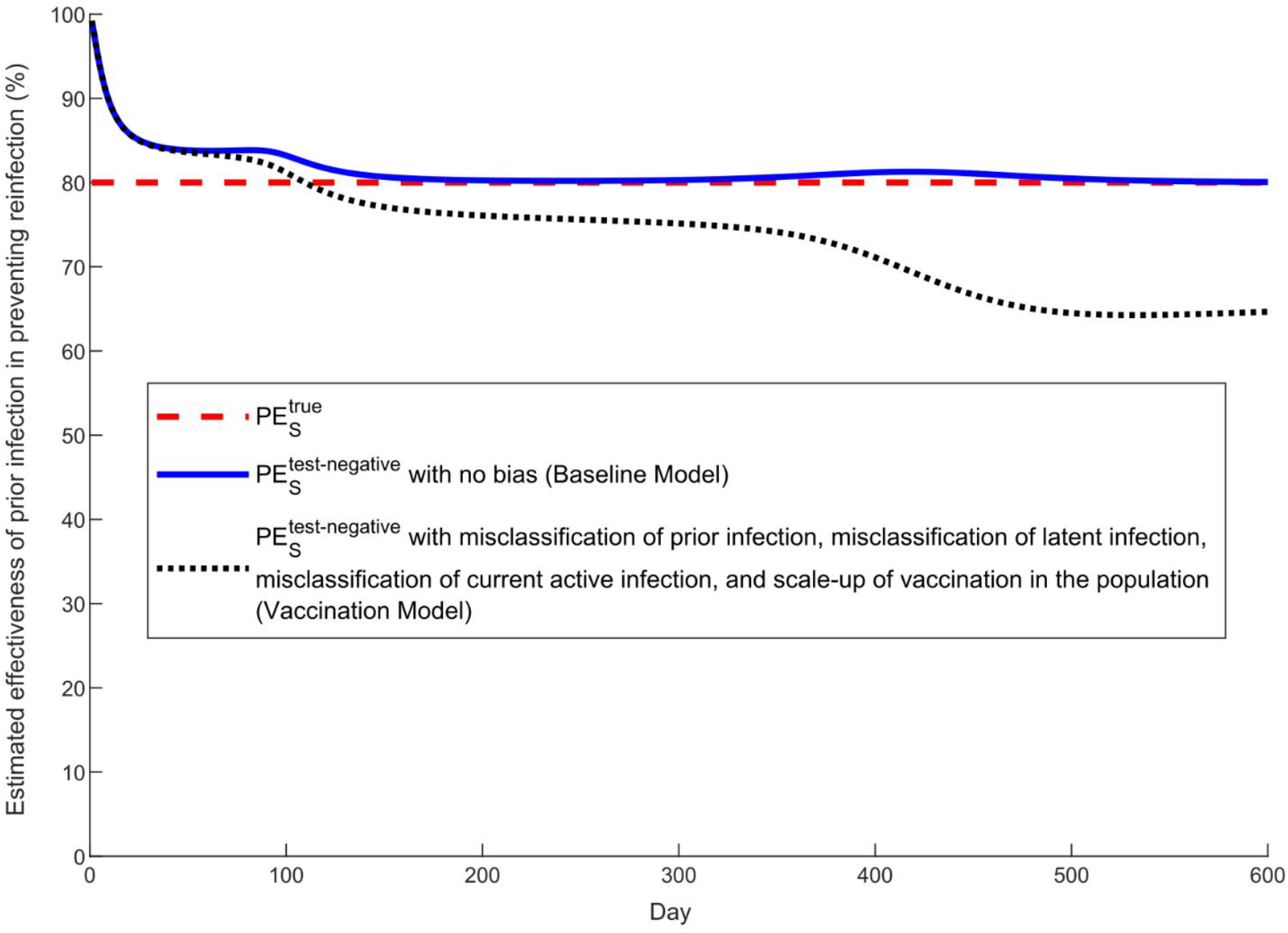
Impact of combined biases in estimating effectiveness of prior infection in preventing reinfection using the test-negative study design 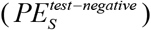. This figure shows the results applying simultaneously misclassification of prior infection, misclassification of latent infection, misclassification of current active infection, and scale-up of vaccination in the population.

## References

1. Abu-Raddad LJ, Chemaitelly H, Malek JA, et al. Assessment of the Risk of Severe Acute Respiratory Syndrome Coronavirus 2 (SARS-CoV-2) Reinfection in an Intense Reexposure Setting. Clin Infect Dis 2021; 73: e1830–e40.

2. Abu-Raddad LJ, Chemaitelly H, Coyle P, et al. SARS-CoV-2 antibody-positivity protects against reinfection for at least seven months with 95% efficacy. EClinicalMedicine 2021; 35: 100861.

3. Abu-Raddad LJ, Chemaitelly H, Ayoub HH, et al. Introduction and expansion of the SARS-CoV-2 B.1.1.7 variant and reinfections in Qatar: A nationally representative cohort study. PLoS Med 2021; 18: e1003879.

4. Chemaitelly H, Bertollini R, Abu-Raddad LJ, National Study Group for C-E. Efficacy of Natural Immunity against SARS-CoV-2 Reinfection with the Beta Variant. N Engl J Med 2021.

5. Hansen CH, Michlmayr D, Gubbels SM, Molbak K, Ethelberg S. Assessment of protection against reinfection with SARS-CoV-2 among 4 million PCR-tested individuals in Denmark in 2020: a population-level observational study. Lancet 2021; 397: 1204–12.

6. Kojima N, Shrestha NK, Klausner JD. A Systematic Review of the Protective Effect of Prior SARS-CoV-2 Infection on Repeat Infection. Eval Health Prof 2021; 44: 327–32.

7. Leidi A, Koegler F, Dumont R, et al. Risk of reinfection after seroconversion to SARS-CoV-2: A population-based propensity-score matched cohort study. Clin Infect Dis 2021: 2021.03.19.21253889.

8. Lumley SF, O’Donnell D, Stoesser NE, et al. Antibody Status and Incidence of SARS-CoV-2 Infection in Health Care Workers. N Engl J Med 2021; 384: 533–40.

9. Pilz S, Chakeri A, Ioannidis JP, et al. SARS-CoV-2 re-infection risk in Austria. Eur J Clin Invest 2021; 51: e13520.

10. Seedat S, Chemaitelly H, Ayoub HH, et al. SARS-CoV-2 infection hospitalization, severity, criticality, and fatality rates in Qatar. Sci Rep 2021; 11: 18182.

11. Angulo FJ, Finelli L, Swerdlow DL. Estimation of US SARS-CoV-2 Infections, Symptomatic Infections, Hospitalizations, and Deaths Using Seroprevalence Surveys. JAMA Netw Open 2021; 4: e2033706.

12. Jones JM, Stone M, Sulaeman H, et al. Estimated US Infection- and Vaccine-Induced SARS-CoV-2 Seroprevalence Based on Blood Donations, July 2020-May 2021. JAMA 2021; 326: 1400–9.

13. Ayoub HH, Chemaitelly H, Seedat S, et al. Mathematical modeling of the SARS-CoV-2 epidemic in Qatar and its impact on the national response to COVID-19. J Glob Health 2021; 11: 05005.

14. Kahn R, Schrag SJ, Verani JR, Lipsitch M. Identifying and alleviating bias due to differential depletion of susceptible people in post-marketing evaluations of COVID-19 vaccines. medRxiv 2021: 2021.07.15.21260595.

15. World Health Organization. Tracking SARS-CoV-2 variants. Available from: https://www.who.int/en/activities/tracking-SARS-CoV-2-variants/. Accessed on: June 5, 2021. 2021.

16. Jackson ML, Nelson JC. The test-negative design for estimating influenza vaccine effectiveness. Vaccine 2013; 31: 2165–8.

17. Verani JR, Baqui AH, Broome CV, et al. Case-control vaccine effectiveness studies: Preparation, design, and enrollment of cases and controls. Vaccine 2017; 35: 3295–302.

18. Lopez Bernal J, Andrews N, Gower C, et al. Effectiveness of Covid-19 Vaccines against the B.1.617.2 (Delta) Variant. N Engl J Med 2021.

19. Sheikh A, McMenamin J, Taylor B, Robertson C. SARS-CoV-2 Delta VOC in Scotland: demographics, risk of hospital admission, and vaccine effectiveness. The Lancet 2021; 397: 2461–2.

20. Nasreen S, He S, Chung H, et al. Effectiveness of COVID-19 vaccines against variants of concern, Canada. medRxiv 2021: 2021.06.28.21259420.

21. Dean NE, Hogan JW, Schnitzer ME. Covid-19 Vaccine Effectiveness and the Test-Negative Design. N Engl J Med 2021.

22. Abu-Raddad LJ, Chemaitelly H, Butt AA, National Study Group for Covid Vaccination. Effectiveness of the BNT162b2 Covid-19 Vaccine against the B.1.1.7 and B.1.351 Variants. N Engl J Med 2021; 385: 187–9.

23. Chemaitelly H, Yassine HM, Benslimane FM, et al. mRNA-1273 COVID-19 vaccine effectiveness against the B.1.1.7 and B.1.351 variants and severe COVID-19 disease in Qatar. Nat Med 2021; 27: 1614–21.

24. National Project of Surveillance for Variants of Concern and Viral Genome Sequencing. Qatar viral genome sequencing data. Data on randomly collected samples. https://www.gisaid.org/phylodynamics/global/nextstrain/. 2021 [cited; Available from: https://www.gisaid.org/phylodynamics/global/nextstrain/

25. Hasan MR, Kalikiri MKR, Mirza F, et al. Real-Time SARS-CoV-2 Genotyping by High-Throughput Multiplex PCR Reveals the Epidemiology of the Variants of Concern in Qatar. Int J Infect Dis 2021; 112: 52–4.

26. Chemaitelly H, Tang P, Hasan MR, et al. Waning of BNT162b2 Vaccine Protection against SARS-CoV-2 Infection in Qatar. N Engl J Med 2021; 385: e83.

27. Benslimane FM, Al Khatib HA, Al-Jamal O, et al. One Year of SARS-CoV-2: Genomic Characterization of COVID-19 Outbreak in Qatar. Front Cell Infect Microbiol 2021; 11: 768883.

28. Tang P, Hasan MR, Chemaitelly H, et al. BNT162b2 and mRNA-1273 COVID-19 vaccine effectiveness against the SARS-CoV-2 Delta variant in Qatar. Nat Med 2021; 27: 2136–43.

29. Ayoub HH, Chemaitelly H, Makhoul M, et al. Epidemiological impact of prioritising SARS-CoV-2 vaccination by antibody status: mathematical modelling analyses. BMJ Innov 2021; 7: 327–36.

30. Ayoub HH, Chemaitelly H, Mumtaz GR, et al. Characterizing key attributes of COVID-19 transmission dynamics in China’s original outbreak: Model-based estimations. Glob Epidemiol 2020; 2: 100042.

31. Abu-Raddad LJ, Chemaitelly H, Ayoub HH, et al. Characterizing the Qatar advanced-phase SARS-CoV-2 epidemic. medRxiv 2020: 2020.07.16.20155317v2 (non-peer-reviewed preprint).

32. Mumtaz GR, El-Jardali F, Jabbour M, Harb A, Abu-Raddad LJ, Makhoul M. Modeling the Impact of COVID-19 Vaccination in Lebanon: A Call to Speed-Up Vaccine Roll Out. Vaccines (Basel) 2021; 9.

33. Makhoul M, Ayoub HH, Chemaitelly H, et al. Epidemiological Impact of SARS-CoV-2 Vaccination: Mathematical Modeling Analyses. Vaccines (Basel) 2020; 8.

34. Makhoul M, Chemaitelly H, Ayoub HH, Seedat S, Abu-Raddad LJ. Epidemiological Differences in the Impact of COVID-19 Vaccination in the United States and China. Vaccines (Basel) 2021; 9: 2021.01.07.21249380.

35. Abu-Raddad LJ, Chemaitelly H, Ayoub HH, et al. Waning of mRNA-1273 vaccine effectiveness against SARS-CoV-2 infection in Qatar. medRxiv 2021: 2021.12.16.21267902.

36. Abu-Raddad LJ, Chemaitelly H, Ayoub HH, et al. Association of Prior SARS-CoV-2 Infection With Risk of Breakthrough Infection Following mRNA Vaccination in Qatar. JAMA 2021; 326: 1930–9.

37. MATLAB®. The Language of Technical Computing. The MathWorks, Inc. 2019.

38. Goldberg Y, Mandel M, Bar-On YM, et al. Protection and waning of natural and hybrid COVID-19 immunity. medRxiv 2021: 2021.12.04.21267114.

39. European Centre for Disease Prevention and Control. Rapid increase of a SARS-CoV-2 variant with multiple spike protein mutations observed in the United Kingdom. Available from: https://www.ecdc.europa.eu/sites/default/files/documents/SARS-CoV-2-variant-multiple-spike-protein-mutations-United-Kingdom.pdf. Accessed on: February 10, 2021. 2020 [cited; Available from:

40. Galloway SE, Paul P, MacCannell DR, et al. Emergence of SARS-CoV-2 B.1.1.7 Lineage - United States, December 29, 2020-January 12, 2021. MMWR Morb Mortal Wkly Rep 2021; 70: 95–9.

41. Challen R, Brooks-Pollock E, Read JM, Dyson L, Tsaneva-Atanasova K, Danon L. Risk of mortality in patients infected with SARS-CoV-2 variant of concern 202012/1: matched cohort study. BMJ 2021; 372: n579.

42. Thermo Fisher Scientific. TaqPath™ COVID-19 CE-IVD RT-PCR Kit instructions for use. Available from: https://assets.thermofisher.com/TFS-Assets/LSG/manuals/MAN0019215_TaqPathCOVID-19_CE-IVD_RT-PCR%20Kit_IFU.pdf. Accessed on December 02, 2020. 2020.

43. Abu-Raddad LJ, Chemaitelly H, Ayoub HH, et al. Characterizing the Qatar advanced-phase SARS-CoV-2 epidemic. Sci Rep 2021; 11: 6233.

44. Coyle PV, Chemaitelly H, Ben Hadj Kacem MA, et al. SARS-CoV-2 seroprevalence in the urban population of Qatar: An analysis of antibody testing on a sample of 112,941 individuals. iScience 2021; 24: 102646.

45. Al-Thani MH, Farag E, Bertollini R, et al. SARS-CoV-2 Infection Is at Herd Immunity in the Majority Segment of the Population of Qatar. Open Forum Infect Dis 2021; 8: ofab221.

46. Jeremijenko A, Chemaitelly H, Ayoub HH, et al. Herd Immunity against Severe Acute Respiratory Syndrome Coronavirus 2 Infection in 10 Communities, Qatar. Emerg Infect Dis 2021; 27: 1343–52.

47. Jacoby P, Kelly H. Is it necessary to adjust for calendar time in a test negative design?: Responding to: Jackson ML, Nelson JC. The test negative design for estimating influenza vaccine effectiveness. Vaccine 2013;31(April (17)):2165-8. Vaccine 2014; 32: 2942.

48. Pearce N. Analysis of matched case-control studies. BMJ 2016; 352: i969.

49. Rothman KJ, Greenland S, Lash TL. Modern epidemiology. 3rd ed. Philadelphia: Wolters Kluwer Health/Lippincott Williams & Wilkins; 2008.

50. StataCorp. Stata Statistical Software: Release 17. College Station, TX: StataCorp LLC. 2021.

51. Planning and Statistics Authority-State of Qatar. Qatar Monthly Statistics. Available from: https://www.psa.gov.qa/en/pages/default.aspx. Accessed on: May 26, 2020. 2020.

52. Ayoub HH, Mumtaz GR, Seedat S, Makhoul M, Chemaitelly H, Abu-Raddad LJ. Estimates of global SARS-CoV-2 infection exposure, infection morbidity, and infection mortality rates in 2020. Glob Epidemiol 2021; 3: 100068.

53. Li R, Pei S, Chen B, et al. Substantial undocumented infection facilitates the rapid dissemination of novel coronavirus (SARS-CoV2). Science 2020.

54. Lauer SA, Grantz KH, Bi Q, et al. The Incubation Period of Coronavirus Disease 2019 (COVID-19) From Publicly Reported Confirmed Cases: Estimation and Application. Ann Intern Med 2020.

55. Zou L, Ruan F, Huang M, et al. SARS-CoV-2 Viral Load in Upper Respiratory Specimens of Infected Patients. N Engl J Med 2020.

56. Rothe C, Schunk M, Sothmann P, et al. Transmission of 2019-nCoV Infection from an Asymptomatic Contact in Germany. N Engl J Med 2020; 382: 970–1.

57. World Health Organization. Report of the WHO-China Joint Mission on Coronavirus Disease 2019 (COVID-19). Available from :https://www.who.int/docs/default-source/coronaviruse/who-china-joint-mission-on-covid-19-final-report.pdf. Accessed on March 10, 2020; 2020.

58. United Nations Department of Economic and Social Affairs Population Dynamics. The 2019 Revision of World Population Prospects. Available from https://population.un.org/wpp/. Accessed on March 1st, 2020. 2020.

